# Poor Readability of COVID-19 Vaccine Information for the General Public: A Lost Opportunity

**DOI:** 10.1101/2021.06.11.21258778

**Authors:** Luke S. Bothun, Scott E. Feeder, Gregory A. Poland

## Abstract

**Background:** All adults in the Unites States now have access to COVID-19 vaccines. During the vaccination process, Emergency Use Authorization (EUA) fact sheets are provided.

**Objective:** To analyze the ease of reading (i.e., readability) of the EUA-approved fact sheets for the vaccines currently available in the United States, the V-Safe adverse event survey script, and the Centers for Disease Control and Prevention (CDC) website on COVID-19 vaccines.

**Design:** We analyzed the readability of Pfizer, Moderna, and Janssen EUA fact sheets, as well as the V-Safe survey script and the vaccine-related information on the CDC website.

**Measurements:** Readability factors include the following: average length of paragraphs, sentences, and words; font size and style; use of passive voice; the Gunning-Fog index; the Flesch Reading Ease index; and the Flesch-Kincaid Grade Level index.

**Results:** Only the V-Safe adverse event survey script met readability standards for adequate comprehension. The mean readability scores of the EUA fact sheets and the CDC website were as follows: Flesch Reading Ease score (mean 44.35); Flesch-Kincaid Grade Level (mean 10.48); and Gunning-Fog index (mean 11.8). These scores indicate that a 10^th^-12^th^ grade-level education is necessary to comprehend these documents.

**Conclusion:** The average person in the United States would have difficulty understanding the information provided in the EUA fact sheets and CDC COVID-19 vaccine website; however, the V-Safe survey was written at an appropriate reading level. To ensure that the public fully understands information regarding COVID-19 vaccines, simplified information material should be developed.

## INTRODUCTION

The 2019 coronavirus disease (COVID-19) is a respiratory illness that has resulted in a global pandemic and the subsequent development of vaccines.^1^ Vaccine information for recipients includes several vaccine information documents, including the Centers for Disease Control and Prevention (CDC) website on COVID-19 vaccines and the vaccine-specific emergency-use authorization (EUA) fact sheet for the Pfizer, Moderna, or Janssen vaccine they are receiving. Subjects are also encouraged to sign up for an adverse event tracking survey called V-Safe. The CDC website on COVID-19 and the EUA fact sheets contain basic vaccine information, including potential benefits, side effects, and necessary precautions. The purpose of this study was to analyze the readability of the EUA fact sheets, the CDC website on COVID-19 information, and the V-Safe adverse event survey script in the United States to determine if they meet guidelines for adequate public readability.

Vaccine recipients should be able to understand vaccine information in order to adequately understand possible risks and benefits of receiving the vaccine. Because these documents are the primary method of communicating vaccine information to the public, the readability of these materials is critical. Determining the readability of such documents is necessary to make sure that the text is written at an adequate comprehension level for the general public with differing reading skills. A National Work Group on Literacy and Health study demonstrated that approximately 25% of U.S. citizens have very low-level reading skills and are unable to comprehend medication instructions or a bus schedule.^2^ The National Adult Literacy Agency suggests that reading materials for the general population should have a 7^th^-grade readability level, which is the average reading level for adults in the United States.^3^ To achieve this, many experts advise using plain wording, active voice, short sentences, and present tense.^4^

## MATERIALS and METHODS

We acquired the following vaccine information documents: COVID-19 vaccine EUA fact sheets, which are given to vaccine recipients in the U.S., from Pfizer, Moderna, and Janssen; the full V-Safe adverse event survey script; and COVID-19 vaccine information found on the CDC website (i.e. Questions and Answers; Does it Work?; Is it safe?; Are there side effects?; and What if I am pregnant or breastfeeding?) for the general public.^5^ Although the length of the V-Safe script varies based on the reported symptoms, we reviewed the entire script.

Two study authors independently analyzed the readability metrics of each document using Microsoft Word readability tools. Formatting metrics, including total length, sentence length, font size and style, and percentage of passive sentences, in addition to three standard readability formulas were used to analyze the reading difficulty of each document.

The Flesch Reading Ease score is a readability formula that predicts ease of reading on a scale from 1 to 100, with higher scores indicating easier readability and 70 being the level of an average U.S. reader.^4^ The Flesch-Kincaid Grade Level score is a readability index that determines the ease of readability on a scale from 1 to 12. This score corresponds to the education-grade level necessary to comprehend a passage.^4^ Table 1 compares the Flesch-Kincaid Grade Level score and Flesch Reading Ease scores. The Gunning-Fog index is a readability index that scores from 1 to 20 based on the number of complex or polysyllabic words in a sentence and the number of sentences in a roughly 100-word passage. The most reliable score is obtained by averaging the Gunning-Fog index score for three random 100-word passages throughout a document. A Gunning-Fog index score of 7 is necessary to achieve near-universal comprehension in the United States (See Table 2).^6^

**TABLE 1:** Flesch Reading Ease Score Comparison ^4^

| Style | Flesch Reading Ease Score | Average Sentence Length in Words | Average No. of Syll. Per 100 Words | Type of Magazine | Estimated School Grade Completed | Estimated Percent of US Adults |
| --- | --- | --- | --- | --- | --- | --- |
| Very Easy | 90-100 | 8 or less | 123 or less | Comics | 4<sup>th</sup> grade | 93 |
| Easy | 80-90 | 11 | 131 | Pulp Fiction | 5<sup>th</sup> grade | 91 |
| Fairly Easy | 70-80 | 14 | 139 | Slick Fiction | 6<sup>th</sup> grade | 88 |
| Standard | 60-70 | 17 | 147 | Digests | 7<sup>th</sup> or 8<sup>th</sup> grade | 83 |
| Fairly Difficult | 50-60 | 21 | 155 | Quality | Some high school | 54 |
| Difficult | 30-50 | 25 | 167 | Academic | High school to college | 33 |
| Very Difficult | 0-30 | 29 or more | 192 or more | Scientific | College | 4.5 |

**TABLE 2:** Gunning-Fog Index Grade Comparison

| Gunning-Fog Index | Reading Level by Grade |
| --- | --- |
| 17 | College Graduate |
| 16 | College Senior |
| 15 | College Junior |
| 14 | College Sophomore |
| 13 | College Freshman |
| 12 | High school Senior |
| 11 | High school Junior |
| 10 | High school Sophomore |
| 9 | High school Freshman |
| 8 | 8<sup>th</sup> Grade |
| 7 | 7<sup>th</sup> Grade |
| 6 | 6<sup>th</sup> Grade |

## RESULTS

We reviewed the COVID-19 EUA vaccine fact sheets for the Pfizer, Moderna, and Janssen vaccines, the complete V-Safe adverse event survey script, and CDC information about COVID-19 vaccines. No significant differences were detected in the scoring metrics between the two independent study authors who analyzed readability.

Document length ranged from 5 to 18 pages (mean 8.25). The number of words in the documents (including headers) ranged from 1,662 to 5,564 (mean 3,086.8). The average words per page ranged from 175.9 to 334.2 (mean 294.8), the average words per paragraph ranged from 7.0 to 25.1 (mean 17.76), and the average word length ranged from 4.9 to 5.4 characters (mean 5.18). The font size ranged from 11 to 16 points, and all documents used sans serif fonts.

The average sentence length ranged from 9.8 to 18.7 words (mean 14.4) (see Table 3), and the percent of passive sentences ranged from 0.2% to 21.1% (mean 15.04%). The Flesch Reading Ease Score ranged from 40.9 to 66.8 (mean 48.84). The Flesch-Kincaid Grade Level ranged from 6.3 to 11.6 (mean 9.76). The Gunning-Fog Score ranged from 8.03 to 12.43 (mean 11.19). See Table 4.

**TABLE 3:** Publicly Accessible COVID-19 Vaccine Document Statistics

|  | Total Pages | Total Paragraphs | Total Words | Words/ Page | Words/ Paragraph | Font Style | Font Size | Word Length (Characters) | Sentence Length (Words) |
| --- | --- | --- | --- | --- | --- | --- | --- | --- | --- |
| V-Safe Survey Script | 28 | 624 | 4,924 | 175.9 | 7.0 | Calibri | 11 | 4.9 | 9.8 |
| Pfizer EUA Fact Sheet | 5 | 101 | 1,671 | 334.2 | 16.5 | Arial | 12 | 5.4 | 14.1 |
| Moderna EUA Fact Sheet | 5 | 83 | 1,662 | 332.4 | 20.0 | Arial | 12 | 5.2 | 15 |
| Janssen EUA Fact Sheet | 5 | 80 | 1,613 | 322.6 | 20.2 | Arial | 12 | 5.3 | 14.4 |
| CDC COVID-19 Vaccine Website | 18 | 222 | 5,564 | 309.1 | 25.1 | Arial | 14-16 | 5.1 | 18.7 |
| Recommended | <15 | None | None | None | None | Arial/Calibri | ≥14 | None | 12-17 |

**TABLE 4:**
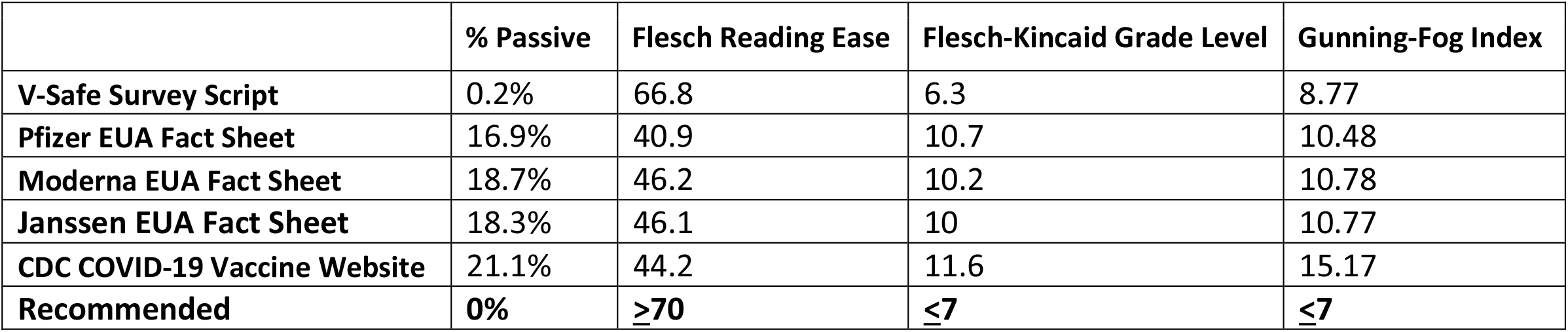
Publicly Accessible COVID-19 Vaccine Document Readability Scores

## CONCLUSIONS

To our knowledge, this is the first study on readability for COVID-19 vaccine information currently used for the general public in the United States. We found that only the V-Safe adverse event survey script met most of the recommended readability levels for average vaccine recipient. The remaining information sources we reviewed did not met readability standards.

Readability studies demonstrate that the general public in the United States is less likely to understand long documents.^7^ Such studies recommend that health information documents should be shorter than 15 pages.^8^ The vaccine information documents in this study had an acceptable page length average of 8.25, but the range extended to 18 pages, with an outlier of 28 pages. High word density also raises the difficulty level of a text; therefore, it is advised that sentences are a maximum of 12 – 17 words long.^9^ Although the average sentence length (14.4 words) in our study was acceptable, the range extended to 18.7 words.

Font style and size also play roles in analyzing the readability of a text. Sans serif fonts (i.e., Arial and Calibri) are easier to read than serif fonts (i.e., Times New Roman), and smaller fonts such as size 12 are harder to read than larger fonts such as size 14.^10^ All documents used an appropriate font style, but the size 12 font found in every document was too small. Many passive-voice sentences, or sentences with the subject receiving the action of the verb, also decrease readability.^4^ The average percentage of passive-voice sentences in our study was fairly low, but the range extended up to 21%.

Because nearly half of adults in the United States read at a 7^th^-grade level,^11^ vaccine information materials provided for the currently available COVID-19 vaccines are too difficult to understand for many adult readers. Since it is important that vaccine recipients are able to comprehend information materials, documents with easier readability should be developed and used.

In addition to simplified readability, the use of instructional graphics and multimedia, including pictures, videos, colors, slideshows, and charts, can also increase comprehension.^12^ These resources were used on the CDC website and the V-Safe survey script but not in the EUA vaccine fact sheets. Further studies are required to determine other cultural and educational barriers to the most useful methods of patient education.

Our study is limited to the use of readability statistics pertaining only to education levels from the United States. The strength of our study includes comparisons between widely used and easily accessible online material regarding COVID-19 vaccines and standardized metrics to assess readability.

In conclusion, our study demonstrates that a significant portion of the U.S. general public would only be able to comprehend the V-Safe script out of the informational documents we studied. However, use of the V-Safe script requires a smart phone. Given the importance of these documents to inform and build trust within the community regarding COVID-19 vaccines, greater effort must be applied to improve the readability of these information documents.

## Data Availability

Available upon request

## ACKNOWLEDGEMENTS

We thank the companies, health care workers, and tens of thousands of vaccine recipients around the world involved in the fight against COVID-19.

## Notes

### Competing Interest Statement

Dr. Poland is the chair of a Safety Evaluation Committee for novel investigational vaccine trials being conducted by Merck Research Laboratories. Dr. Poland provides consultative advice on vaccine development to Merck & Co., Medicago, GlaxoSmithKline, Sanofi Pasteur, Johnson & Johnson/Janssen Global Services LLC, Emergent Biosolutions, Dynavax, Genentech, Eli Lilly and Company, Kentucky Bioprocessing, Bavarian Nordic, AstraZeneca, Exelixis, Regeneron, Janssen, Vyriad, Moderna, and Genevant Sciences, Inc. These activities have been reviewed by the Mayo Clinic Conflict of Interest Review Board and are conducted in compliance with Mayo Clinic Conflict of Interest policies. Dr. Poland holds patents related to vaccinia, influenza, and measles peptide vaccines. Dr. Poland has received grant funding from ICW Ventures for preclinical studies on a peptide-based COVID-19 vaccine. This research has been reviewed by the Mayo Clinic Conflict of Interest Review Board and was conducted in compliance with Mayo Clinic Conflict of Interest policies. The remaining two authors, Luke S. Bothun, Scott E. Feeder, M.S., do not have any disclosures.

### Funding Statement

Not funded

